# United States Provider Experiences with Telemedicine for Hepatitis C Treatment: A Nationwide Survey

**DOI:** 10.1101/2024.05.12.24307239

**Authors:** Pruthvi Patel, Martin T. Wells, Elaine Wethington, Martin Shapiro, Yasir Parvez, Shashi N Kapadia, Andrew H Talal

## Abstract

**Background:** Hepatitis C virus (HCV) elimination requires treatment access expansion, especially for underserved populations. Telehealth has the potential to improve HCV treatment access, although data are limited on its incorporation into standard clinical practice.

**Methods:** We conducted a cross-sectional, e-mail survey of 598 US HCV treatment providers who had valid email addresses and 1) were located in urban areas and had written ≥20 prescriptions for HCV treatment to US Medicare beneficiaries in 2019-20 or 2) were located in non-urban areas and wrote any HCV prescriptions in 2019-20. Through email, we notified providers of a self-administered electronic 28-item survey of clinical strategies and attitudes about telemedicine for HCV.

**Results:** We received 86 responses (14% response rate), of which 75 used telemedicine for HCV in 2022. Of those 75, 24% were gastroenterologists/hepatologists, 23% general medicine, 17% infectious diseases, and 32% non-physicians. Most (82%) referred patients to commercial laboratories, and 85% had medications delivered directly to patients. Overwhelmingly, respondents (92%) felt that telehealth increases healthcare access, and 76% reported that it promotes or is neutral for treatment completion. Factors believed to be “extremely” or “very” important for telehealth use included patient access to technology (86%); patients’ internet access (74%); laboratory access (76%); reimbursement for video visits (74%) and audio-only visits (66%). Non-physician licensing and liability statutes were rated “extremely” or “very” important by 43% and 44%, respectively.

**Conclusions:** Providers felt that telehealth increases HCV treatment access. Major limitations were technological requirements, reimbursement, and access to ancillary services. These findings support the importance of digital equity and literacy to achieve HCV elimination goals.

## Background

The hepatitis C virus (HCV) epidemic parallels the ongoing rise in injection drug use.^1,2^ In the United States, persons who use drugs (PWUD) account for the fastest growing population with incident and prevalent HCV infections.^2^ Following the introduction of the first direct acting antivirals (DAAs) in 2013, there was rapid development of treatments which were efficacious and well tolerated.^3–5^ The efficacy of these treatments led the World Health Organization (WHO) and United States Department of Health and Human Services (HHS) to establish the goal of achieving HCV elimination by 2030.^6,7^ A decade later, we are far from attaining this goal ^8,9^, as the majority of the seventy million people living with HCV worldwide remain untreated. Among the greatest remaining HCV elimination obstacles is insufficient access to the highly curative DAAs by the often-underserved populations most affected.^10^

Barriers to healthcare access occur at the patient, provider, and system levels.^11^ Lack of access to HCV treatment often results from the scarcity of HCV-treating providers in a geographic region, or to the logistical difficulties, including stigma, that patients face when accessing specialized health systems.^12,13^ Telehealth, because of its ability to overcome temporal and geographical obstacles, may be a means of improving treatment access for some underserved populations.^14^

The effective use of telehealth to treat HCV has been described in clinical trials or small demonstration programs and a recently completed large randomized controlled trial.^15^ Three controlled trials comparing teleconsultation to face-to-face treatment also demonstrated no difference in sustained virological response (SVR).^16–18^ Expanding on this concept, pilot studies have shown increased engagement and dramatically improved HCV cure rates when telemedicine was provided at opioid treatment programs (OTPs).^19^ The use of telemedicine has been well-received by patients, including people who use drugs (PWUD).^15,20^ Expansion of telemedicine may play an important role in facilitating HCV treatment, and thereby helping to support elimination goals.

The uptake of telemedicine rapidly increased during the COVID-19 pandemic, initially as an approach to minimize infection risk by reducing in-person interactions. Although there were significant decreases in HCV screening and linkage-to-care during the height of COVID-related “lockdown” restrictions ^21^, there have also been a number of HCV telemedicine demonstration projects during the pandemic that illustrate its potential as a tool for HCV treatment.^22–26^ The use of this technology in standard clinical practice, especially when applied to underserved populations, however, is not yet well-understood. In particular, clinicians may need to adapt their usual practices for telemedicine, in order to obtain laboratory testing or imaging. To address this knowledge gap, we conducted a survey to understand how US HCV treatment providers have incorporated telemedicine as a real-world strategy for HCV treatment coinciding with the COVID-19 pandemic.

## Methods

### Design and approval

We conducted a cross-sectional survey among HCV treatment providers across the United States who endorsed using telemedicine for treating HCV in 2022. The study was approved by the Biomedical Research Alliance of New York (BRANY) Institutional Review Board.

### Study population and sampling

To identify HCV treatment providers, we used the “Medicare Part D Providers by Provider and Drug” database to identify National Provider Identifier (NPI) numbers associated with prescriptions for HCV DAAs in 2019 and 2020 (the latest available data at the time of our search in Spring 2023). From these 1,954 providers, we selected providers whose taxonomy codes indicated a specialty of Gastroenterology, Infectious Diseases, Internal Medicine, Family Medicine, Nurse Practitioner, or Physician Assistant (n=1883) as these were considered the specialties most likely to provide HCV care. We then restricted the sample to 1) providers located in urban areas providers who had written ≥20 prescriptions for DAAs in 2019-20, or 2) nonurban providers who had written any prescription for DAAs in 2019-20. Urbanicity was determined by the Rural Urban Commuting Area 2-category coding system.^27^ These interventions resulted in 918 providers from the Medicare database. We linked these NPI numbers to the CarePrecise Healthcare Provider Database, a commercially available dataset that contains provider specialty and contact information.^28^ Of the 918, 734 providers could be linked to valid e-mail addresses and were included in our survey distribution.

### Data Collection

The 28-question survey instrument was designed by 4 investigators with expertise in HCV treatment and survey design (PP, SK, EW, AT). Survey questions focused on providers’ experiences using telemedicine, provider practices specific to HCV treatment through telemedicine, and healthcare workers’ attitudes about telemedicine for HCV treatment (Appendix 1). The survey was pilot-tested among 3 physicians and a social psychologist with experience treating HCV using telemedicine before deployment. Survey invitations were distributed by email from June to September 2023. Participants who were eligible for the full survey received an $50 electronic gift card, and those eligible for a partial survey (*i.e*., those who started the survey but screened out because they did not use telemedicine for HCV treatment) received a $10 electronic gift card.

### Analysis

We performed descriptive analyses of all variables. We compared telemedicine practices and attitudes based on provider specialty, provider practice setting, and the volume of HCV patients managed, using Chi-square or Fisher’s exact test as appropriate. We applied bootstrap subsampling to assess the robustness of statistical estimates, which involves repeatedly sampling observations from the original dataset with replacement to create multiple bootstrap samples. These are reported as bootstrap confidence-intervals alongside survey responses. Statistical analyses were performed using STATA v18.

## Results

We distributed the survey to 734 email addresses of which 136 were invalid, resulting in 598 valid recipients. We received 86 responses (14% response rate), of which 75 endorsed using telemedicine for HCV in 2022 and thus completed the full survey. Characteristics of these 75 participants, including their volume of HCV telemedicine visits and video visits, are shown in Table 1. Providers perceived high efficacy of treatment via telemedicine. Of the 67 providers who reported prescribing HCV treatment at telemedicine visits, 59 (88%) reported that more than 75% of those patients completed treatment, and 72 (97%) reported that more than 75% of those patients achieved SVR12.

**Table 1:**
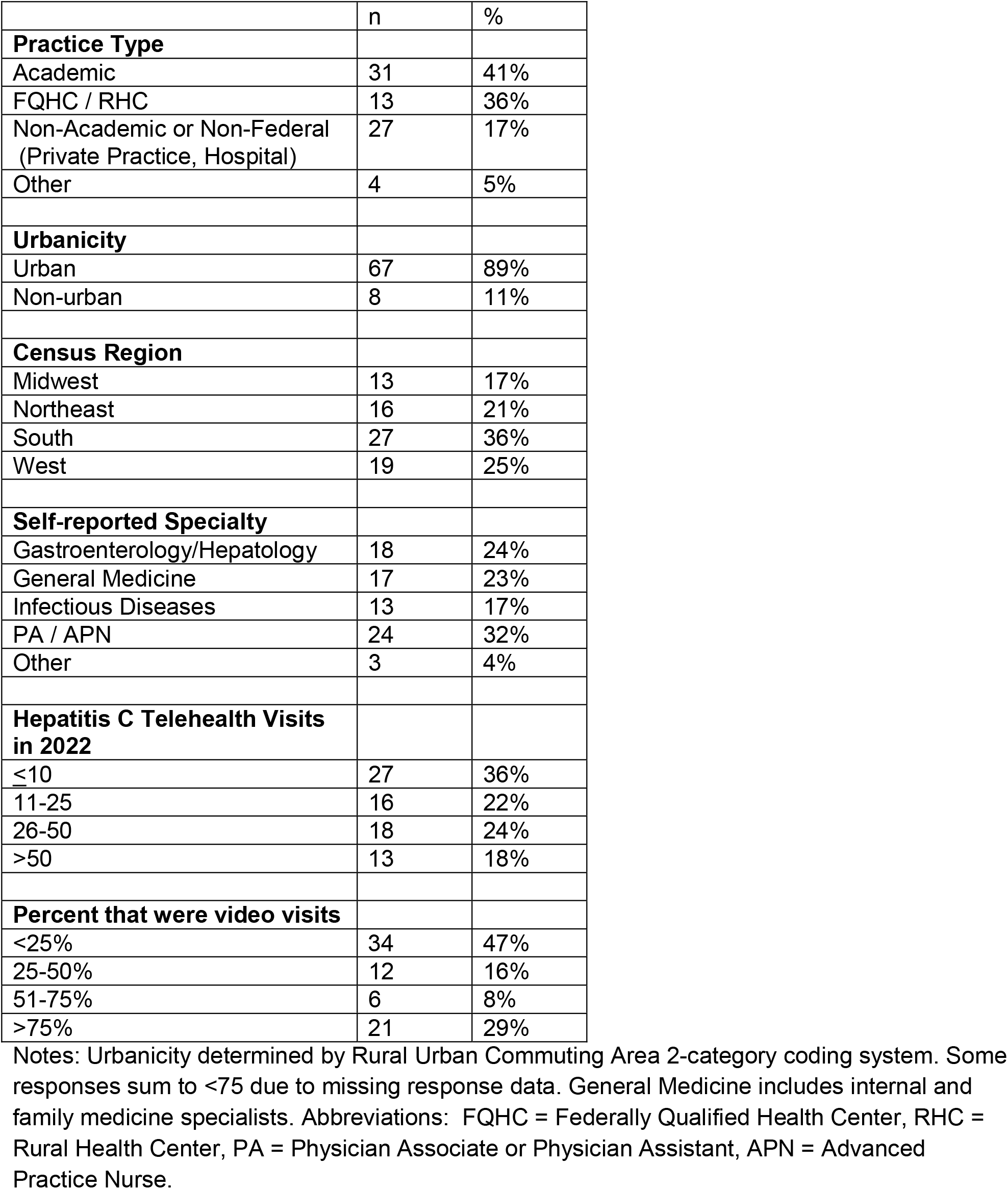
Description of Sample.

### Telehealth clinical practices

Providers reported how they conducted typical steps in HCV management for patients seen by telehealth (Table 2). Most reported that they referred patients to an external laboratory for blood work (82%), to an external radiology facility for imaging (70%), and had prescriptions mailed directly to patients (85%) - implying that many patients could complete treatment without visiting the providers’ office even once. In fewer cases, providers conducted lab work or imaging at their offices (46% and 55% of providers, respectively). Most providers used either the electronic health record software or a dedicated telehealth software program; a smaller number reported using an enterprise/business software platform, such as Zoom or WebEx.

**Table 2.**
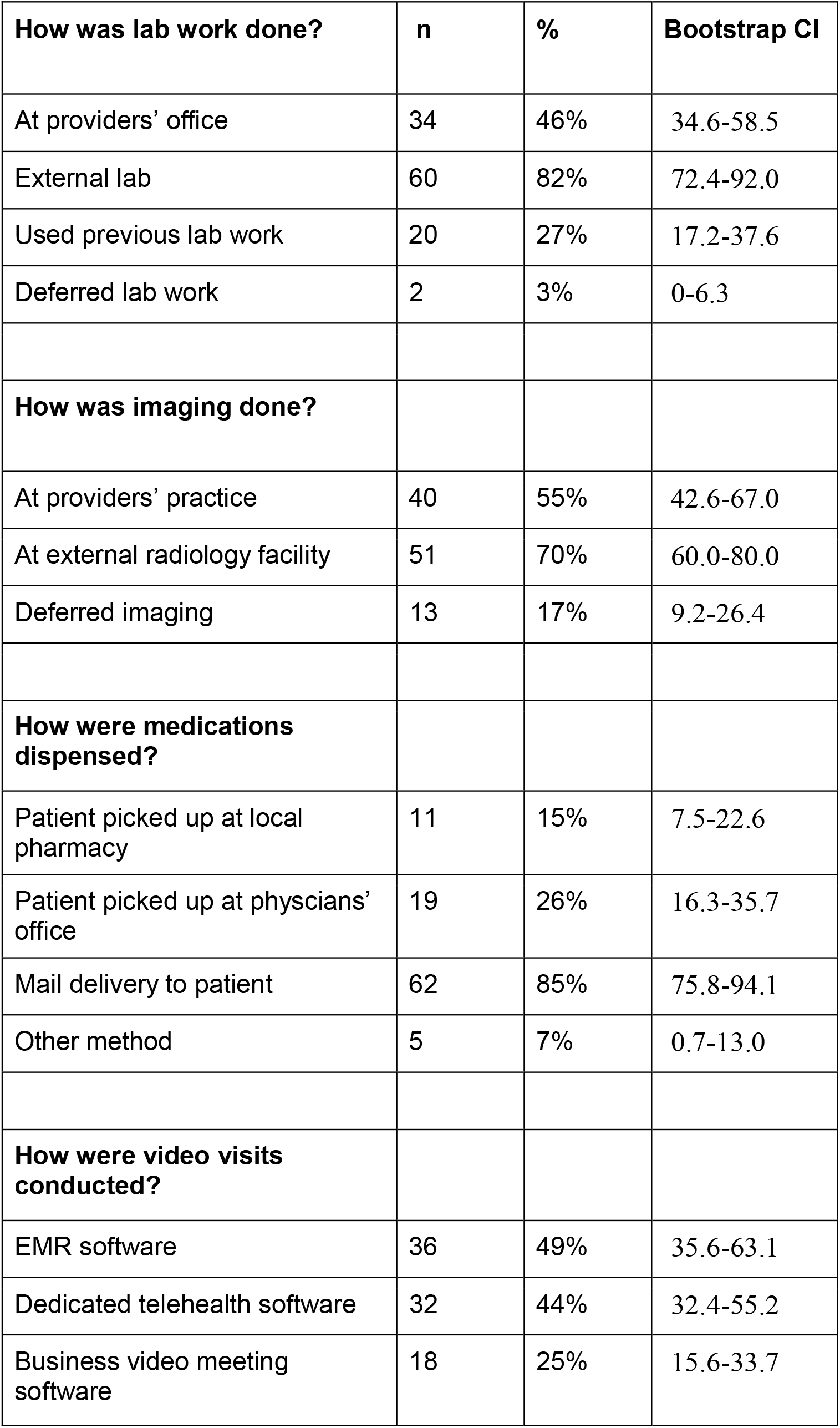
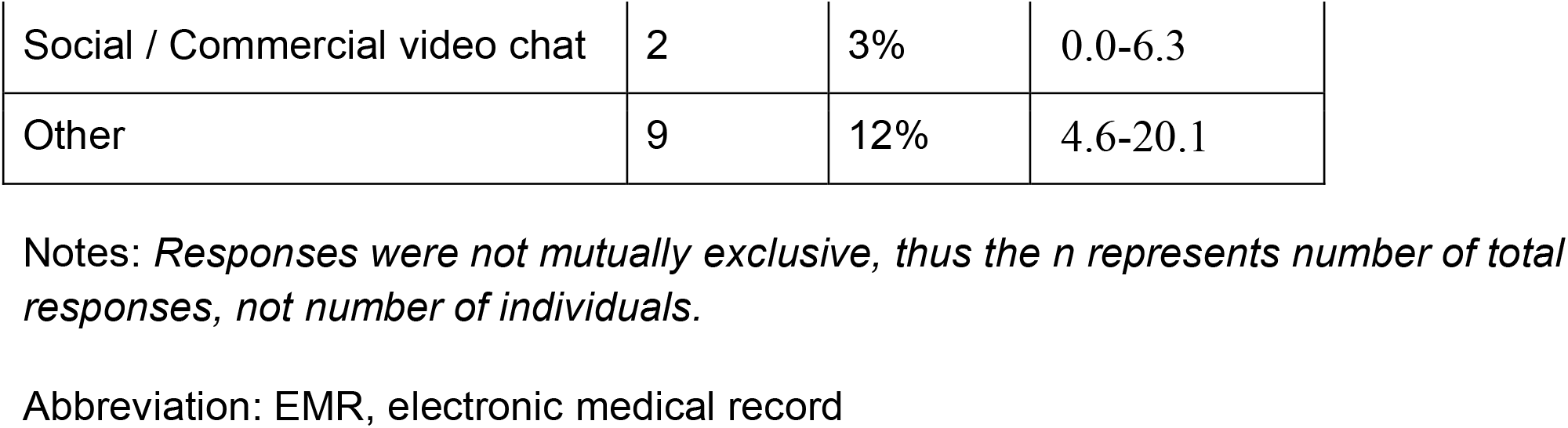
Clinical Practices.

| **How was lab work done?** | **n** | **%** | **Bootstrap CI** |
| --- | --- | --- | --- |
| At providers' office | 34 | 46% | 34.6-58.5 |
| External lab | 60 | 82% | 72.4-92.0 |
| Used previous lab work | 20 | 27% | 17.2-37.6 |
| Deferred lab work | 2 | 3% | 0-6.3 |
| **How was imaging done?** |  |  |  |
| At providers' practice | 40 | 55% | 42.6-67.0 |
| At external radiology facility | 51 | 70% | 60.0-80.0 |
| Deferred imaging | 13 | 17% | 9.2-26.4 |
| **How were medications dispensed?** |  |  |  |
| Patient picked up at local pharmacy | 11 | 15% | 7.5-22.6 |
| Patient picked up at physicians' office | 19 | 26% | 16.3-35.7 |
| Mail delivery to patient | 62 | 85% | 75.8-94.1 |
| Other method | 5 | 7% | 0.7-13.0 |
| **How were video visits conducted?** |  |  |  |
| EMR software | 36 | 49% | 35.6-63.1 |
| Dedicated telehealth software | 32 | 44% | 32.4-55.2 |
| Business video meeting software | 18 | 25% | 15.6-33.7 |
| Social / Commercial video chat | 2 | 3% | 0.0-6.3 |
| Other | 9 | 12% | 4.6-20.1 |

Notes: *Responses were not mutually exclusive, thus the n represents number of total responses, not number of individuals.*
Abbreviation: EMR, electronic medical record

### Telehealth Attitudes

Regarding provider attitudes to telehealth (Table 3), a large majority (92%) of respondents believed it increased access to care, and 76% indicated that it either promoted treatment completion or had no effect on completion compared to in-person visits. Few respondents reported patient concerns about privacy. Most respondents indicated that telehealth was more time-efficient for the provider. When asked about patient engagement, 42/75 (56%) expressed patient engagement concerns: 17% mentioned the inability to perform a physical exam as a concern, 16% reported patient technological literacy, 16% reported patient access to the internet, and 7% respondents expressed difficulty with patient rapport (Appendix Table 1).

**Table 3:** Telehealth Attitudes.

| **How does telehealth affect access to care?** | **n** | **%** | **Bootstrap CI** |
| --- | --- | --- | --- |
| Increases | 67 | 92% | 83.3-96.2 |
| No change | 2 | 3% | 1.2-6.2 |
| Decreases | 3 | 1% | 0.3-5.7 |
| Not sure / no answer | 3 | 4% | 1.6-12.6 |
| **How does telehealth affect HCV treatment completion?** |  |  |  |
| Promotes treatment completion | 31 | 42% | 32.0-53.6 |
| No effect | 25 | 34% | 24.7-45.3 |
| Makes it more difficult | 6 | 8% | 3.8-16.8 |
| Not sure / no answer | 11 | 15% | 8.4-25.5 |
| **Did patients have privacy concerns?** |  |  |  |
| Yes, many patients | 1 | 1% | 0.2-6.4 |
| Yes, but only a few | 4 | 5% | 2.3-12.4 |
| No | 68 | 93% | 86.4-96.7 |
| Not sure / no answer | 0 | 0% | -- |
| **Is telehealth more time-efficient for the provider?** |  |  |  |
| Yes | 40 | 55% | 44.2-65.0 |
| No | 26 | 36% | 25.4-47.3 |
| Not sure / no answer | 7 | 10% | 4.4-19.5 |

### Factors informing the continued use of telehealth

We asked providers to rate the importance of each of several factors on their decision to continue using telehealth for providing care to HCV patients (Figure 1). The factors most often rated “extremely” or “very” important were patients’ having access to computers or mobile devices (86%), patients’ access to laboratories (76%), and patients’ access to the internet (74%). Similarly important were reimbursement for video visits (74%) and audio only phone visits (66%). In contrast, non-physician licensing and liability statutes were seen as less important, rated “extremely” or “very” important by 43% and 44% respectively.

**Figure 1.**
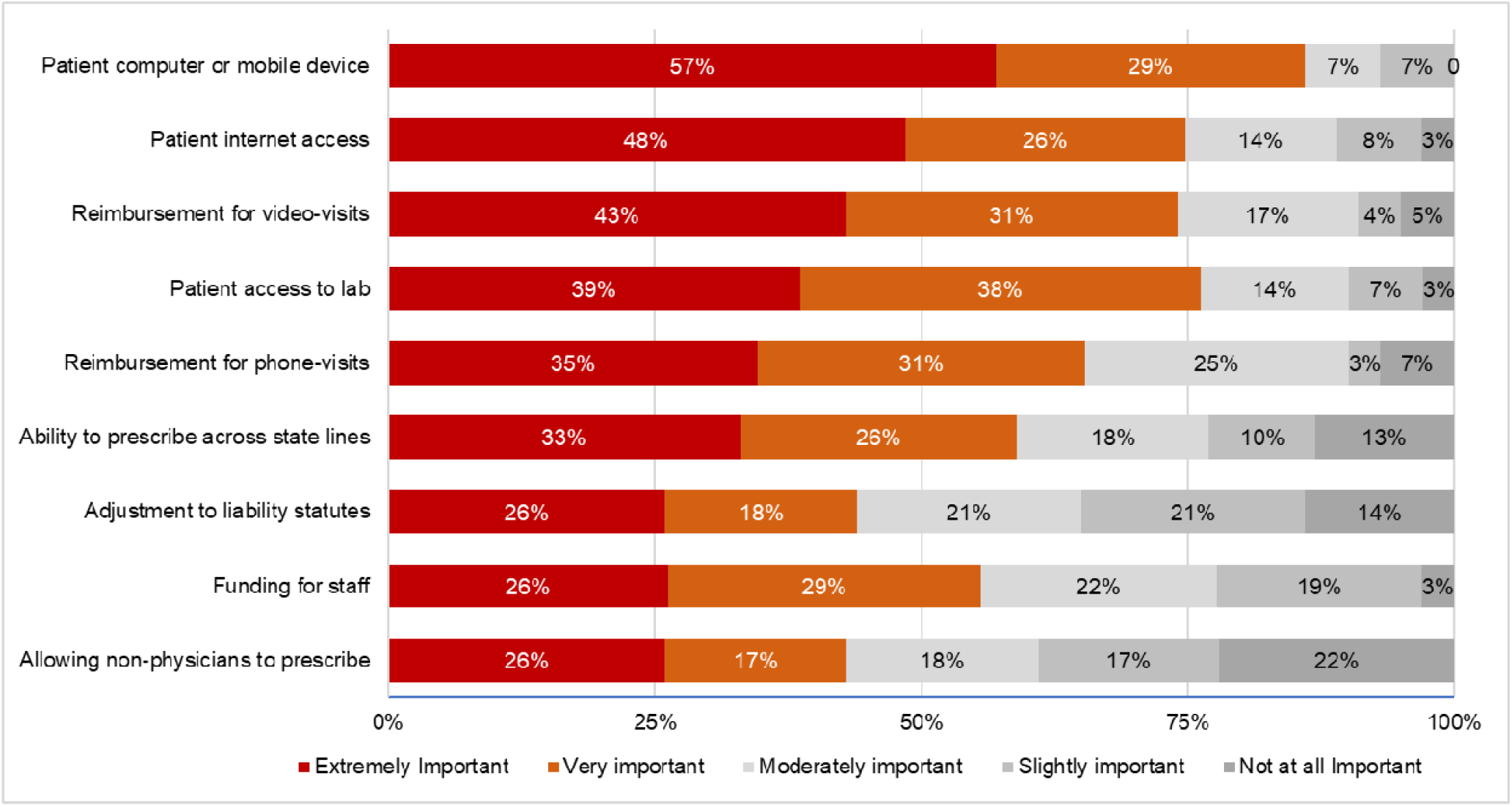
Important factors for continued use of telehealth.

## Discussion

This real-world survey of HCV providers describes high effectiveness of telemedicine-based treatment and its important role in the care paradigm. Over three-quarters of patients completed therapy and were successful in achieving an HCV cure. Although half the respondents reported some concern over patient engagement, they did not feel completion rates using telemedicine were worse than in-person care. The vast majority (92%) of respondents felt that telemedicine improved access to care.

Though the utility of telemedicine is valued by survey respondents, they overwhelmingly stated that patient access to mobile devices or computer and internet connections were the most important barriers for continued use of telemedicine. Health equity in telehealth, as defined by the US Department of Health and Human Services (HSS),^29^ requires making changes in digital literacy, technology, and analytics to enable everyone to receive the health care they need and deserve, regardless of social or economic status. Providing access to fundamental digital infrastructure is needed to expand telemedicine to reach underserved and difficult-to-reach populations. Until infrastructure improvements occur, payment parity of audio-only visits may play an important role to increase healthcare access. Of note, over two-thirds of respondents to this survey reported the importance of payment parity for future telehealth use. A study conducted in New York City in 2020 showed that primary care providers in high-Social Vulnerability Index areas were twice as likely to use telephones as their primary telehealth modality instead of video.^30^ Other attributes of telemedicine, including privacy, as well as funding for additional staff or non-physician prescribers were deemed to be less important barriers for continued use by the respondents.

Most providers reported using external facilities for labs and imaging, and direct mail shipments for medication. The flexibility of these approaches allow patients to fully avoid travel for face-to-face visits during treatment, although it requires that external laboratory facilities are accessible, which may not be the case for all patients. The MinMon study demonstrated that minimal monitoring can still result in high SVR rates for patients treated for hepatitis C.^31^ In the trial, there was no pre-treatment genotyping and the entire DAA course was dispensed on the initial treatment day with no visits or laboratory monitoring until the SVR assessment 12 weeks post-treatment completion. During the study, there were only two telephone check-ins. The authors reported that 89% of participants showed 100% adherence and 95% achieved an SVR. This study suggests that for many HCV-infected patients, successful treatment can be achieved with minimal monitoring, which is often limited to pre-treatment workup and confirmation of cure. These approaches reduce patient travel burden.

There have been several models proposed for utilizing telehealth to expand HCV treatment. One of the earliest projects, ECHO, used telementoring for HCV treatment. The project began in 2011 by Arora *et al*. in which specialists telementored primary care providers to treat HCV in remote areas. The outcome of this long-term project showed no difference in SVR percentages for individuals treated directly by specialists compared to primary care physicians, although the study was conducted in the interferon era.^32^ Another approach has assessed facilitated telemedicine integrated into OTPs, thereby providing access to technology and internet in a “safe” environment for the PWUD population. Project TEAM-C was a randomized trial of 12 OTP sites where telemedicine was integrated into the facility. In this trial, participants felt the facilitated experience provided a safe space for engagement and treatment, with 90.3% achieving SVR in facilitated telemedicine arm compared to 39.4% in referral.^15,33^ These percentages are very similar to that obtained (93%) in a single arm pilot study. ^19^ These studies, along with the opinions reported by our survey population, highlight the promise that telemedicine holds for attaining and achieving national and international goals of HCV elimination.

The strengths of this study include the diversity of respondents’ clinical specialties, which has as similar distribution as national studies of HCV treating providers,^34^ and the geographic regions represented. There has been a recent push to increase access to HCV treatment by expanding the provider pool beyond specialty care.^35,36^ In this survey, a quarter of respondents were primary care physicians and a third were nurse practitioners or physician assistants. Therefore, our sample is reflective of the recent changes in the disciplines of HCV treatment prescribers. Limitations of this investigation include the modest response rate and sample size, particularly for rurally-located providers, despite our attempts to oversample from this group. The sample was selected to be representative of many HCV-treating providers, but based on our sampling scheme, it more likely to represent providers who participate in Medicare and those who only treat a moderate or high volume of HCV patients. An additional limitation, given our sampling approach, is the potential that some HCV providers who had used telemedicine prior to the COVID-19 pandemic may have discontinued their use after the pandemic (*i.e*., in 2022).

## Conclusion

Participants in this survey report the positive role of telemedicine in expanding access to HCV treatment. At the same time, digital equity, payor-parity, and clarifying the best practices for laboratory and imaging, will all be important for the continued use and expansion of this modality to achieve WHO and HHS goals of HCV elimination by 2030.

## Supporting information

Supplement - Survey

## Data Availability

All data produced in the present study are available upon reasonable request to the authors

