## Supplemental Table 1 for "United States Provider Experiences with Telemedicine for Hepatitis C Treatment: A Nationwide Survey"

**Appendix Table 1:**

**Patient Engagement Concerns:**

| | | |
|---|---|---|
| Not as thorough / no exam / labs | 13 | 23.64% |
| Patient technical literacy | 12 | 21.82% |
| Access to tech / internet | 12 | 21.82% |
| Rapport | 5 | 9.09% |
| Technical issues | 4 | 7.27% |
| Loss to follow up | 3 | 5.45% |
| Privacy | 2 | 3.64% |
| Patient distracted | 2 | 3.64% |
| Waiting time | 1 | 1.82% |
| Patient education | 1 | 1.82% |
| **Total** | 55 | 100.00% |

Notes: *Response categories are derived from free-text responses. Responses were not mutually exclusive, thus the n represents number of total responses, not number of individuals.*
