## Supplement - Survey for "United States Provider Experiences with Telemedicine for Hepatitis C Treatment: A Nationwide Survey"

Survey Research Institute

AT CORNELL UNIVERSITY

### HCV Telehealth Treatment Survey

Thank you for taking the time to complete our survey. The purpose of this survey is to understand how hepatitis C treatment providers are incorporating telemedicine into their practices, particularly as a strategy for HCV elimination.

You can refuse to answer any questions that you want, and you can stop the survey at any time. We expect the survey will take about 15-20 minutes to complete. Your responses will remain confidential outside of the study team.

This study has been reviewed by the BRANY Institutional Review Board of New York. For specific questions, you may contact Dr. Andrew Talal (the lead investigator of this survey) at.

You can find additional information about the study and download here.

Next

HCV Telehealth Treatment Survey

Have you prescribed hepatitis C treatment for at least one patient since January 1 2022?

- Yes
- No

Previous Next
Finish Later

Progress 0% 20% 40% 60% 80% 100%

**The first few questions will ask about your clinical practice. If you work in multiple practices, please enter the responses that describe where you see the majority of your patients with Hepatitis C.**

Where is your principal practice located? (Enter US Zip Code)

Which of the following best describes your practice type?

- ○ Private Practice
- ○ Academic Medical Center
- ○ Federally Qualified Healthcare Center (FQHC)
- ○ Opioid Treatment Program (i.e. Methadone clinic)
- ○ Veterans Affairs
- ○ Other - Please specify: ______

About how many unique Hepatitis C patients did you see in 2022?

- ○ 0-50
- ○ 51-100
- ○ 101-200
- ○ More than 200

Previous | Next

Finish Later

Progress 0% 20% 40% 60% 80% 100%

Have you EVER used telehealth (video or telephone encounters in lieu of an in-person visit) to manage Hepatitis C positive patients?

- ○ Yes
- ○ No

If you have never used telehealth to evaluate and/or treat patients with Hepatitis C, what were the reason(s)? (Select all that apply).

- ☐ Reimbursement too low
- ☐ Difficulty with ascertainment of labs and/or imaging
- ☐ Did not have the infrastructure (i.e. software)
- ☐ Did not have the support staff
- ☐ Significant portion of patients did not have access to technology or internet
- ☐ Did not feel it provided the same quality of care as in-person visits
- ☐ Institution did not support use
- ☐ Licensing, credentialing or other restrictions
- ☐ My patients are not interested in using telehealth
- ☐ I do not have the time to incorporate this into my practice
- ☐ Prior authorization for HCV meds require in-person visit
- ☐ I have liability or malpractice concerns
- ☐ Other reasons - Please specify: ____________

Previous | Next

Finish Later

HCV Telehealth Treatment Survey

Progress 0% 20% 40% 60% 80% 100%

In calendar year 2022, did you have any telehealth encounters (video or telephone encounters in lieu of in-person visit) for Hepatitis C positive patients?

- ○ Yes
- ○ No

If you did not use telehealth for managing hepatitis C in 2022, what were the reasons? (Select all that apply).

- ☐ Used telehealth only during COVID-19 related "lockdown"
- ☐ Stopped using telehealth because of lower reimbursement
- ☐ Stopped using telehealth because the technology did not work well
- ☐ Patients no longer requested telehealth encounters
- ☐ Stopped using telehealth because of malpractice concerns
- ☐ Stopped using telehealth because of change in practice environment (e.g. practice stopped supporting it, or you moved to a practice that did not support it)
- ☐ Stopped using telehealth because of inability to conduct laboratory or radiology tests
- ☐ Stopped using telehealth because of inability to conduct physical exams
- ☐ Prior authorization for HCV meds require in-person visit
- ☐ Other reasons - Please specify: ______

Previous | Next

Finish Later

Progress 0% 20% 40% 60% 80% 100%

In the year 2022, approximately how many outpatient Hepatitis C consultations (initial or follow-up) did you conduct using telehealth?

- Less than or equal to 10
- 11 to 25
- 26 to 50
- Greater than 51

In the year 2022, for approximately how many of your patients did you prescribe anti-viral therapy for Hepatitis C during telehealth visits?

- None
- 1-20
- 21-50
- More than 51

What percentage of these patients completed treatment?

- Less than 25%
- 25% - 50%
- 51% - 75%
- More than 75%
- Do not know

What percentage of the patients who started treatment achieved sustained virologic response (SVR) 12 weeks post-treatment?

- Less than 25%
- 25% - 50%
- 51% - 75%
- More than 75%
- Do not know

Previous Next
Finish Later

Progress 0% 20% 40% 60% 80% 100%

For patients you saw using telehealth in 2022, how did you conduct lab work? (Select all that apply)

- ☐ Patients came to provider's office
- ☐ Patients went to an external lab
- ☐ Used previous lab results as a guide
- ☐ Deferred lab work

For patients you saw using telehealth in 2022, how was imaging done? (Select all that apply)

- ☐ Conducted in our practice setting (ie clinic or hospital)
- ☐ Patients went to an external radiology facility
- ☐ Deferred imaging

For patients you saw using telehealth in 2022, how did you deliver medications? (Select all that apply)

- ☐ Patient picked up at local pharmacy
- ☐ Patients came to the office to pick up
- ☐ Mail delivery to the patient's residence
- ☐ Other - Please specify: ______

Previous | Next

Finish Later

Progress: 0% 20% 40% 60% 80% 100%

**The term "telehealth" can include both audio and video visits.**

In 2022, what percentage of your telehealth encounters for hepatitis C were video-visits?

- ◯ Less than 25%
- ◯ 25% - 50%
- ◯ 51% - 75%
- ◯ More than 75%

If you used a video service, which of the following did you use to conduct the telehealth visit? (Select all that apply)

- ☐ Telehealth functionality directly integrated into electronic health record software (i.e. Epic)
- ☐ Dedicated telehealth platform (i.e. Doxy-me, Doximity)
- ☐ Enterprise/Business video meeting software (i.e. Zoom, Webex)
- ☐ Commercial social video-chat software (i.e. WhatsApp, Facetime, Google Meet)
- ☐ Other - Please specify: ______

Previous | Next

Finish Later

Progress
0% 20% 40% 60% 80% 100%

In your opinion, how does telehealth affect access to care?

- Increases access to care
- No change in access to care
- Decreases access to care
- Not sure

Please describe your patient engagement concerns related to telehealth visits, if any:

Previous Next
Finish Later

HCV Telehealth Treatment Survey

Progress 0% 20% 40% 60% 80% 100%

In your opinion, how does telehealth affect completion of Hepatitis C treatment compared to all in-person visits?

- Promotes treatment completion
- Makes treatment completion more difficult
- No effect
- Not sure

Did Hepatitis C patients express concerns about privacy, security or confidentiality issues using telehealth? (i.e., not having a private location in which to conduct telehealth evaluations)

- Yes, many patients had these concerns
- Yes, but only a few patients had these concerns
- No
- Not sure

Are telehealth visits more efficient with regards to time? (i.e., ability to see more patients during the same clinic session)

- Yes
- No
- Not sure

Previous Next
Finish Later

HCV Telehealth Treatment Survey

Progress

0% 20% 40% 60% 80% 100%

Please rate the importance of each factor on your decision to continue using telehealth in the future for providing care for Hepatitis C patients

| | Extremely important | Very important | Moderately important | Slightly important | Not at all important |
|---|---|---|---|---|---|
| **Reimbursement for video-visits at same level as in-office visits** | ○ | ○ | ○ | ○ | ○ |
| **Reimbursement for telephone-only visits at same level as in-office visits** | ○ | ○ | ○ | ○ | ○ |
| **Patient access to laboratory** | ○ | ○ | ○ | ○ | ○ |
| **Patient internet access** | ○ | ○ | ○ | ○ | ○ |
| **Patient computer or mobile device access** | ○ | ○ | ○ | ○ | ○ |
| **Adjustment to liability statutes** | ○ | ○ | ○ | ○ | ○ |
| **Ability to prescribe across state lines** | ○ | ○ | ○ | ○ | ○ |
| **Funding for staff to aid with remote visits** | ○ | ○ | ○ | ○ | ○ |
| **Allowing non-physicians to prescribe via telehealth** | ○ | ○ | ○ | ○ | ○ |

Previous Next

Finish Later

Progress

0% 20% 40% 60% 80% 100%

Are there any other ideas, concerns, or perspectives about telemedicine for HCV that you would like to share with us?

Previous Next

Finish Later

Progress 0% 20% 40% 60% 80% 100%

**Thank you. For completing the full survey, you will receive a $50 gift card electronic gift card.**

What is your preferred name and email address for delivery of an electronic gift card?

First name:

Last name:

E-mail Address:

May we contact you by email for additional questions or opportunities to participate in research?

○ Yes ○ No

We are interested in surveying as many providers as possible who have perspectives about prescribing HCV treatment using telehealth, including physicians and advanced practice professionals. Would you provide the name and email address of any other providers who you would recommend to complete this survey?

Previous | Submit Survey

Finish Later
